# Designing Efficient Contact Tracing Through Risk-Based Quarantining

**DOI:** 10.1101/2020.11.16.20227389

**Authors:** Andrew Perrault, Marie Charpignon, Jonathan Gruber, Milind Tambe, Maimuna S. Majumder

**Affiliations:** Center for Research on Computation and Society, Harvard University, Cambridge, MA; Institute for Data, Systems, and Society, MIT, Cambridge, MA; Department of Economics, MIT, Cambridge, MA; Computational Health Informatics Program, Boston Children’s Hospital, Boston, MA, USA; Department of Pediatrics, Harvard Medical School, Boston, MA, USA

## Abstract

Contact tracing for COVID-19 is especially challenging because transmission often occurs in the absence of symptoms and because a purported 20% of cases cause 80% of infections, resulting in a small risk of infection for some contacts and a high risk for others. Here, we introduce risk-based quarantine, a system for contact tracing where each cluster (a group of individuals with a common source of exposure) is observed for symptoms when tracing begins, and clusters that do not display them are released from quarantine. We show that, under our assumptions, risk-based quarantine reduces the amount of quarantine time served by more than 30%, while achieving a reduction in transmission similar to standard contact tracing policies where all contacts are quarantined for two weeks. We compare our proposed risk-based quarantine approach against test-driven release policies, which fail to achieve a comparable level of transmission reduction due to the inability of tests to detect exposed people who are not yet infectious but will eventually become so. Additionally, test-based release policies are expensive, limiting their effectiveness in low-resource environments, whereas the costs imposed by risk-based quarantine are primarily in terms of labor and organization.

## 1 Introduction

As cases of coronavirus disease 2019 (COVID-19) pass 28 million worldwide (Dong, Du, and Gardner 2020), a clear distinction has emerged between countries. There are those nations, such as South Korea, that have a very effective regime structured around testing and contact tracing. Aggressive use of both has kept case counts low, minimized damage, and crushed ongoing outbreaks as they emerge. Other nations, most notably the United States (U.S.), have been much less organized in both their testing and contact tracing regimes. As a result, case counts have been high, and transmission of SARS-CoV-2—the virus that causes COVID-19—has been frequent and ongoing.

Clearly, one of the major problems facing the U.S. is a lack of a coherent national testing strategy (Schneider 2020). But another problem is a general failure to establish successful contact tracing, which has suffered from low response rates and a large number of index cases that report having no contacts (Clark, Chiao, and Amirian 2020; Khazan 2020). Investigations regarding the determinants of quarantine adherence and contact reporting behavior suggest that perceived effectiveness and cost of such behaviors are major drivers of participation (Smith et al. 2020; Webster et al. 2020). SARS-CoV-2 faces challenges on both fronts. A typical non-household close contact has a seemingly low probability (less than 6%) of becoming infected (Kucharski et al. 2020; Laxminarayan et al. 2020), most infections are not serious, and because as much as 56% of transmission occurs without symptoms (Casey et al. 2020; Ferretti et al. 2020), infectious individuals may not notice when they transmit the virus to others. Quarantining in the U.S. is also expensive, especially given that a large number of workers do not receive full compensation for time missed from work due to illness (U.S. Department of Labor and U.S. Bureau of Labor Statistics 2019). Combining these factors yields a scenario where isolation (i.e., the separation of infected individuals from others) and quarantine (i.e., the separation of exposed individuals from others) times are perceived to be long and expensive relative to the benefit they provide, resulting in low participation.

In the U.S., a monolithic approach to contact tracing may exacerbate these issues; individuals who are close contacts of known infectious individuals are generally advised to quarantine irrespective of their odds of infection— despite the fact that the probability of transmission varies widely for different contact events. Notably, studies suggest that 80% of infections may be caused by only 10-20% of those who are infected with SARS-CoV-2 (Adam et al. 2020; Endo et al. 2020; Laxminarayan et al. 2020), as is often the case with infectious diseases (Lloyd-Smith et al. 2005). As a result, the effectiveness of quarantine may vary widely depending on the infected individual who is the source of the exposure. To the best of our knowledge, transmission heterogeneity has not been exploited by previous contact tracing systems, with the exception of Kennedy-Shaffer, Baym, and Hanage (2020), who rely on inexpensive, low sensitivity tests to test all contacts for infection.

Of course, while some individuals have a much higher propensity to transmit (e.g., severely ill individuals in the case of the related MERS-CoV (Majumder et al. 2017)), the underlying propensity of any given person is unknown. Contract tracers can, however, observe a strong correlate of this propensity: the number of contacts of a given infected person who develop symptoms. As this number increases, the risk of infection increases for any other individual in the cluster. We argue for differential handling of contacts based on this observation, reserving more severe control measures, such as quarantine, for clusters where symptoms have been observed. We call our system risk-based quarantine (RBQ).

In this paper, we evaluate the potential of RBQ using an agent-based branching process model of SARS-CoV-2 transmission (Kretzschmar et al. 2020; Okolie and Müller 2020; Klinkenberg, Fraser, and Heesterbeek 2006). We embed contact tracing and quarantine into this model. We then introduce an RBQ regime, where clusters are observed for symptoms for a day after being reached by contact tracers and the cluster is released from quarantine if no symptoms are observed.^1^ Such an approach has two advantages; first, individuals spend less time in quarantine, which reduces the costs of this approach, and second, individuals who are in a cluster where there is a reported infection may be more likely to comply with quarantine because there is a clear signal that they are at risk. While our model was developed in the context of the current SARS-CoV-2 epidemic, taking into account the transmission dynamics of the virus known to date and testing processes currently in place, RBQ is also applicable to other infectious diseases with substantial transmission heterogeneity and transmission in the absence of symptoms.

We compare our RBQ regime to test-driven approaches to reducing quarantine, such as releasing individuals from quarantine who test negative. If tests were freely available, had no cost, and could predict with 100% accuracy whether a contact has contracted or will develop SARS-CoV-2 infection, this would clearly be a better approach than requiring individuals to quarantine; however, none of these three conditions can thus far be met in reality (Woloshin, Patel, and Kesselheim 2020; Ramdas, Darzi, and Jain 2020; Wang et al. 2020; Ai et al. 2020; Service 2020; Waltz 2020). To compare RBQ against more realistic testing-based policies, we incorporate testing, including false negatives and delays in processing results, into our model and measure both the associated accuracy and cost. Much attention has been paid to inexpensive, rapid, tests, which may alleviate test performance issues through volume of testing (Guglielmi 2020; Larremore et al. 2020). Nevertheless, these tests are not yet available and implementing rapid testing at national scale is a significant logistical challenge.

## 2 Modeling transmission and superspreading of SARS-CoV-2

To analyze the costs and effectiveness of various contact tracing policies, we use data from a number of U.S. and non-U.S. sources (Table 1) to construct an agent-based branching process model of SARS-CoV-2 transmission over three generations (Figure 1, detailed description in Appendix A). In each simulation, the index case (first generation) represents an individual whose information has been provided to contact tracers. They are assumed to have age drawn from the age distribution of the U.S. population (Howden and Meyer 2011), COVID-19 symptoms that led them to isolate, and a positive test result. The other timing parameters of their infection and transmission are drawn from the distributions specified in Table 1, including the onset of infectiousness, symptoms, and isolation. Age is used in the model in two places: for the purposes of contact generation and to calculate the probability of death given infection.

**Table 1:**
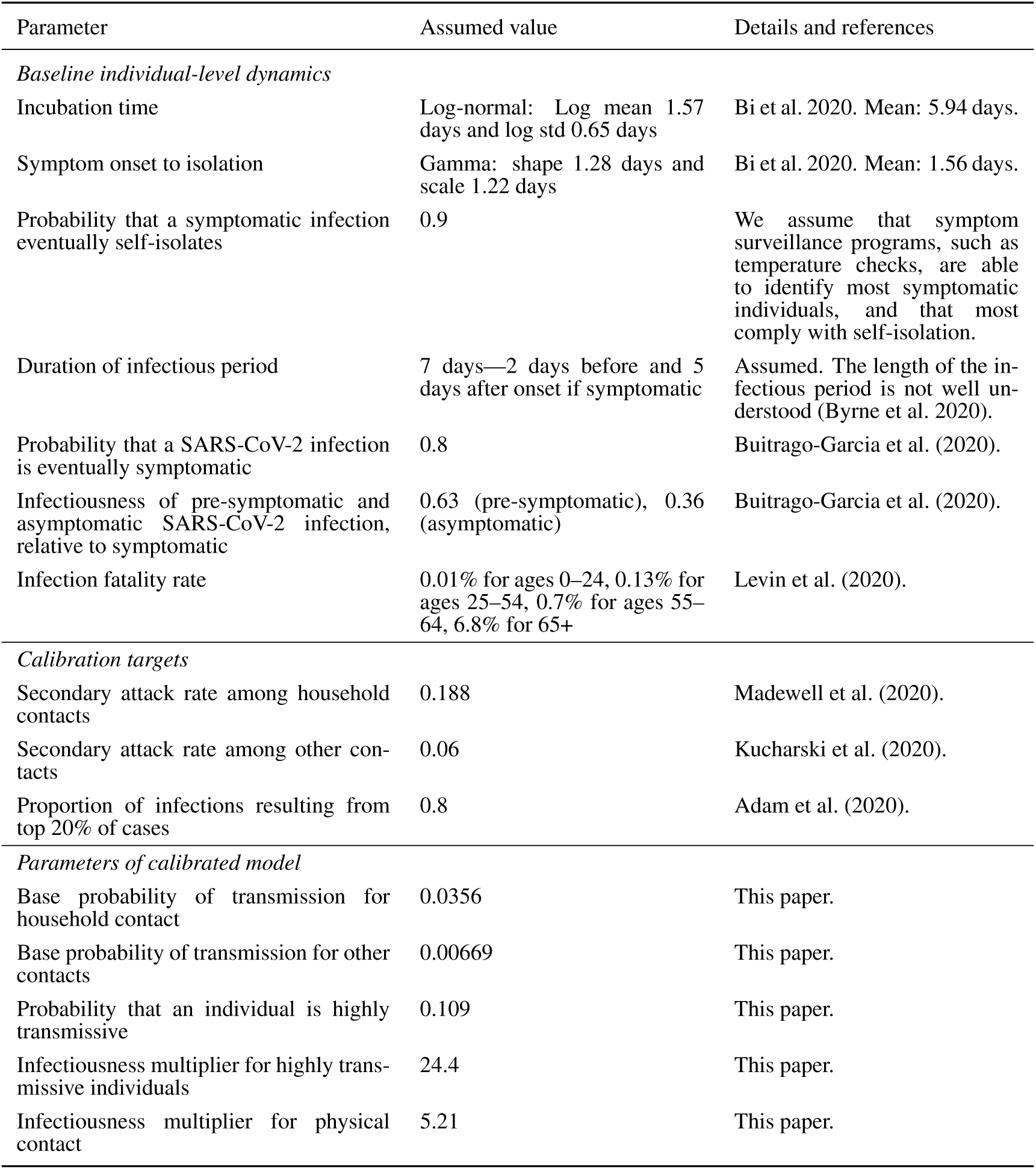
Parameters of the infectiousness model

**Figure 1:**
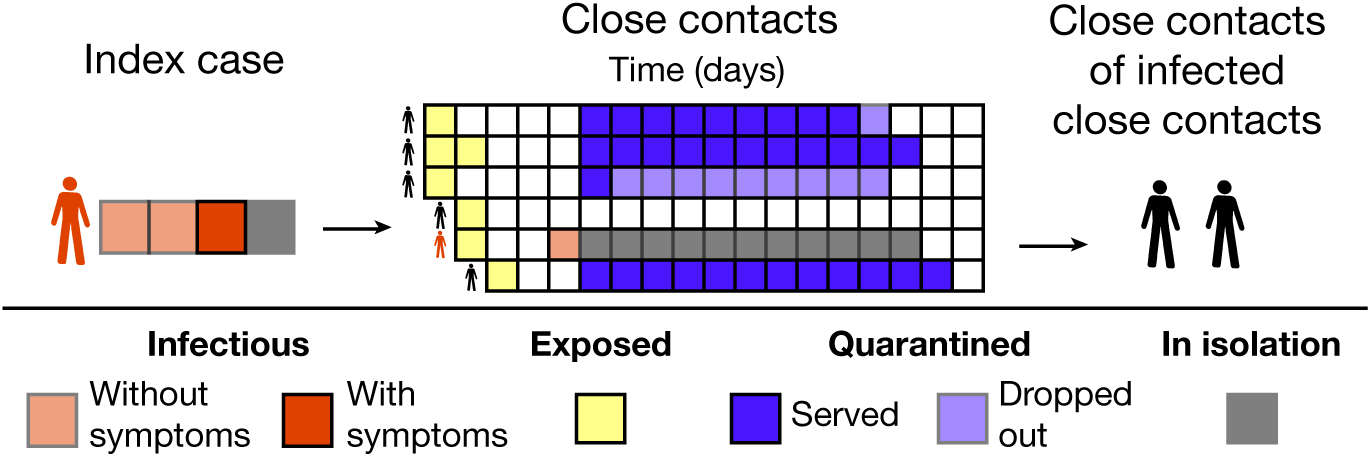
An agent-based branching process model is used to capture the interplay of stochastic behavioral dynamics and contact tracing decision-making. The diagram depicts standard contact tracing with a two-week quarantine period for contacts for an example index case with six contacts during their three-day pre-isolation infectious period.

To generate contacts, the index case is randomly assigned to a respondent from the POLYMOD contact survey (Mossong et al. 2008) with the same age category. This second generation consists of the contacts reported by that respondent, repeated for each day of the index case’s pre-isolation infectious period. Each contact that was reported as daily in POLYMOD is assumed to occur on every day of the pre-isolation infectious period, and contacts that were reported as non-daily are assumed to represent unique individuals. We eliminate contacts with duration of less than 15 minutes, as they are not elicited by contact tracers under CDC guidelines (Centers for Disease Control and Prevention 2020).

Contact tracers have the opportunity to alter the behavior of second generation contacts in order to reduce transmission to their own contacts (i.e., the third generation). We measure the effectiveness of a contact tracing policy through the number of infections it prevents, relative to the amount of transmission that would occur in the absence of control measures. The costs of a contact tracing policy include the days of quarantine served by the second generation contacts and the monetary cost of performing tracing, monitoring and testing.

Figure 1 depicts the application of standard contact tracing with a quarantine period of two weeks from the time of last exposure. The illustrative index case is infectious for two days without symptoms and one day with symptoms before entering isolation. The index case has six contacts: two on day one of infectiousness, two on day two, one on day three, and one on both day one and day two. Because processing the test results takes a day and contact tracing takes a day (see Table 2 for our contact tracing modeling assumptions), the contacts are not asked to quarantine until day 7. All contacts start their quarantine at the same time, but those who had a later last exposure are asked to quarantine for longer. Contact #3 serves only a single day of quarantine before dropping out. Nevertheless, this has no effect on transmission because they are not infectious. Contact #4 cannot be reached by tracers, but also never becomes infectious. At the time of notification, Contact #5 has already become infectious and has exposed two people. They develop symptoms the day after they are asked to quarantine and isolate for ten days after symptom onset. All of the time they spends in quarantine is recorded by the simulation as isolation as we do not consider isolating infectious individuals to have a social cost. The two individuals exposed by Contact #5 form the third generation of the model, but neither becomes infected.

**Table 2:**
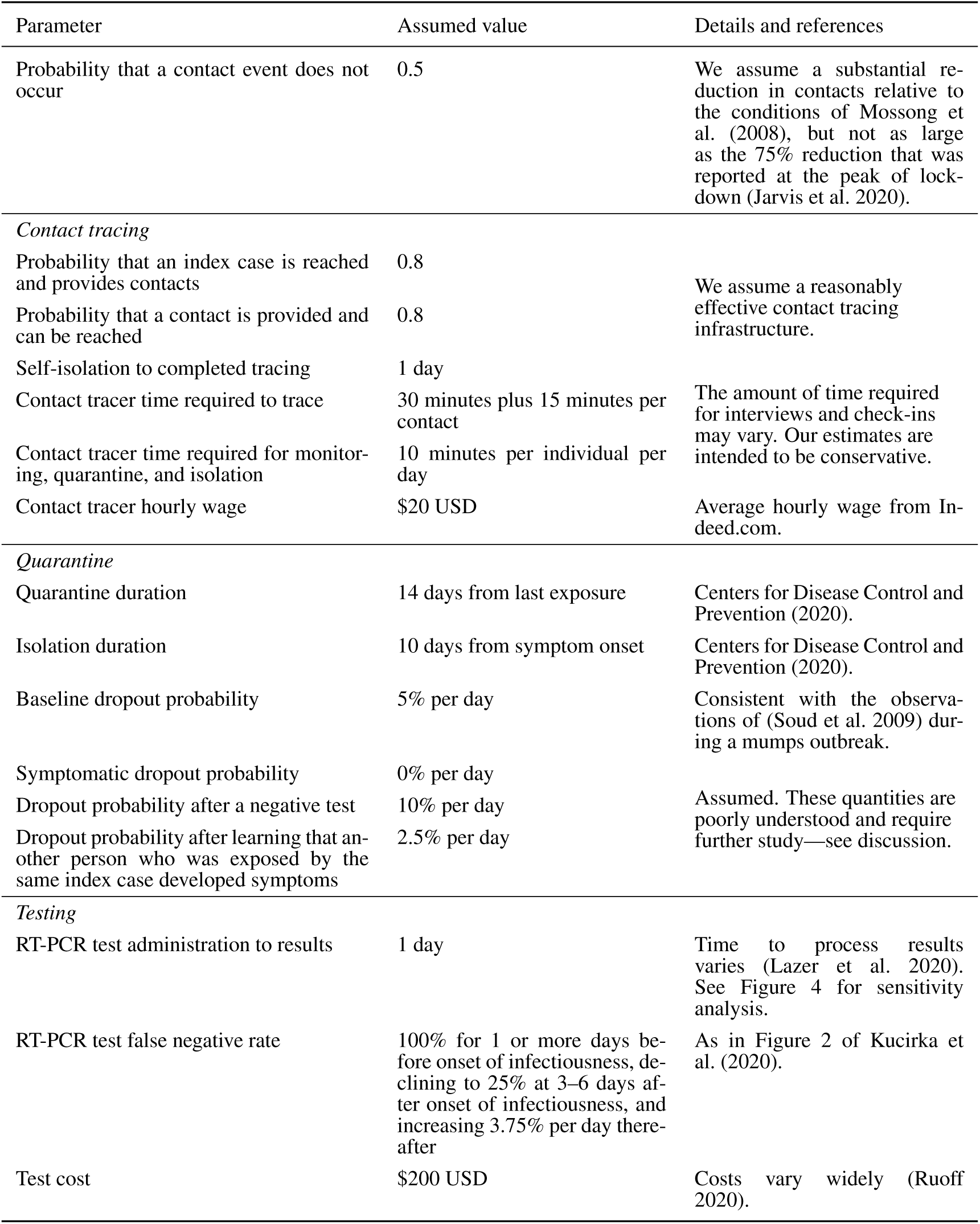
Parameters of the intervention models

### Modeling transmission heterogeneity

Studies on SARS-CoV-2 transmission suggest substantial transmission heterogeneity; 10–20% of cases are likely responsible for 80% of transmission (Adam et al. 2020; Endo et al. 2020; Laxminarayan et al. 2020). To achieve a similar concentration of secondary infections, our model includes three sources of transmission heterogeneity that correspond to hypothesized causes of superspreading events (Shen et al. 2004; Frieden and Lee 2020). First, there is natural variation in the number of close contacts reported by each respondent in POLYMOD, i.e., the top 20% of respondents report 50% of the close contacts. Second, household and physically-close contacts are more likely to result in transmission. Third, each individual has a chance of being highly transmissive, which represents a combination of behavioral and biological factors such as viral shedding and mask wearing. We calculate the probability that a close contact results in transmission as the product of a base transmission probability (which depends on whether the contact occurred within a household or not), a multiplier if the contact was physical, and a multiplier if the transmitter was highly transmissive:

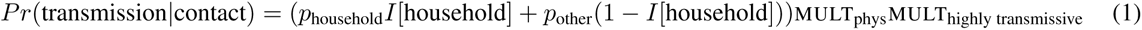

Using Nelder-Mead (Nelder and Mead 1965), we calibrate the values of the parameters p_household_, *p*_other_, MULT_phys_, MULT_highly transmissive_ and *Pr*(highly transmissive) to match the calibration targets of an 18.8% secondary attack rate among household close contacts (Madewell et al. 2020), a 6% attack rate among other close contacts (Kucharski et al. 2020), and such that the top 20% of cases (i.e., the 20% that cause the most infections) cause 80% of infections (Adam et al. 2020).

The calibrated values are reported in Table 1. The resulting model matches the calibration targets exactly and has an *R*_0_ of 1.88, somewhat lower than current estimates (Liu et al. 2020). This lower *R*_0_ is consistent with the fact that most contact tracing studies observe a low *R*_eff_, which may be caused by interventions such as physical distancing or by the difficulty in detecting all transmissions. Of all secondary infections, 17% are caused by asymptomatic cases (i.e., cases that never develop symptoms) and 37% by pre-symptomatic (i.e., cases that will eventually develop symptoms, but have not at time of transmission), broadly consistent with the estimates of 10% and 46% by Ferretti et al. 2020. Of the 20% of cases that result in the most secondary infections, 55% are highly transmissive; they also have 8.7% more close contacts than average (26.3 vs. 24.2), 23% more household contacts (3.62 vs. 2.95), and 4.4% more physical contacts (17.9 vs. 17.2).

There are potentially multiple calibrations matching the targets, because they do not sufficiently distinguish different causes of transmission heterogeneity, e.g., whether physical contact or individual variation in infectiousness are more impactful. Additionally, we do not account for heterogeneous susceptibility across contacts, e.g., due to immunocompromised status. In the discussion, we describe how our results depend on the cause of transmission heterogeneity.

## 3 Using infection risk estimates to drive quarantine policies

A consequence of transmission heterogeneity is that the probability that an exposed individual becomes infected can vary dramatically based on the source and the type of contact event. For example, in our model, a household contact of a highly transmissive individual has nearly a 90% chance of becoming infected. Ideally, we would like contact tracing policies to select different outcomes for contacts that depend on the probability of infection, e.g., enforce a strict quarantine on contacts of highly transmissive individuals and monitor other contacts for symptoms via an app-based check-in. Differential handling of contacts by risk level would, in principle, allow for achieving more transmission reduction at a lower cost in total quarantine days.

Formally, let *s* be the source individual, *e* be the exposed individual, and *c* be the parameters of the close contact. We can calculate the posterior probability that *s* infects *e* by the contact *c*:

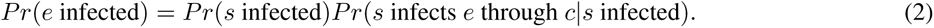

However, we often lack information about the level of infectiousness of the source individual. Highly transmissive individuals are usually only observed retrospectively because it is very difficult to proactively estimate an individual’s level of transmissiveness (Wong et al. 2015). As it appears that individual-level differences in transmissiveness (both behavioral and biological) are a major driver of transmission heterogeneity, Equation 2 may not provide useful information in practice.

We instead propose to retroactively identify highly transmissive individuals through the observed outcomes experienced by close contacts. Intuitively, the longer we monitor the close contacts of a source individual without observing symptoms, the less likely it is that the source individual is highly transmissive. Let *HI*(*s*) denote the event that *s* is highly transmissive. Given symptom observations ***o*** for all close contacts, we can calculate the posterior probability that *s* is highly transmissive using Bayes’ rule:

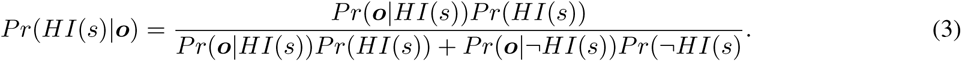

This estimate of the infectiousness level of the source can be inserted into Equation 2 to improve our estimate that *e* is infected and thus drive a differential quarantine policy. We provide a numerical illustration of the combination of Equations 2 and 3 at the end of this section.

### Risk-based quarantine

Equations 2 and 3 can be used to design complex contact tracing policies that exploit all of the available data and the entire space of decisions. We show that a simple policy we call “risk-based quarantine” (RBQ) can yield contact tracing policies that are very effective at reducing transmission while requiring less quarantine than standard quarantine-only policies. RBQ quarantines and observes the close contacts for *n* days. On day *n* + 1, if no close contacts have developed symptoms, they are released into monitoring. The parameter *n* can be selected according to local conditions or depend on cluster size, as larger clusters provide more independent observations of infectiousness. We focus our attention on the case of *n* = 1, where contacts serve a single day of quarantine while they are observed for symptoms before the decision of whether to release the cluster is made.

Figure 2 shows an example of risk-based quarantine with *n* =1 days of observation for a single index case, who is labeled *A* in the figure. Subject *A* has two household contacts, *B* and *C*, and two non-household contacts, *D* and *E*, on days 1–3. On days 1 and 2, *A* is infectious, but pre-symptomatic. On day 3, *A* develops symptoms, and on day 4, *A* self-isolates and is tested (recall that we assume that all index cases self-isolate, seek testing, and remain isolated until they are no longer infectious). On day 5, *A* receives a positive test result and their contacts are asked to quarantine on day 6. In standard contact tracing, these contacts would serve two weeks of quarantine from the day of exposure. Under RBQ, they are released after a day because none of *B*–*E* display symptoms. In the simplest form of RBQ, contact tracers do not remain in contact with *B*–*E* after this point. In the next section, we describe more conservative variants of RBQ that have different conditions for releasing asymptomatic clusters or that continue to monitor *B*–*E* after the end of quarantine.

**Figure 2:**
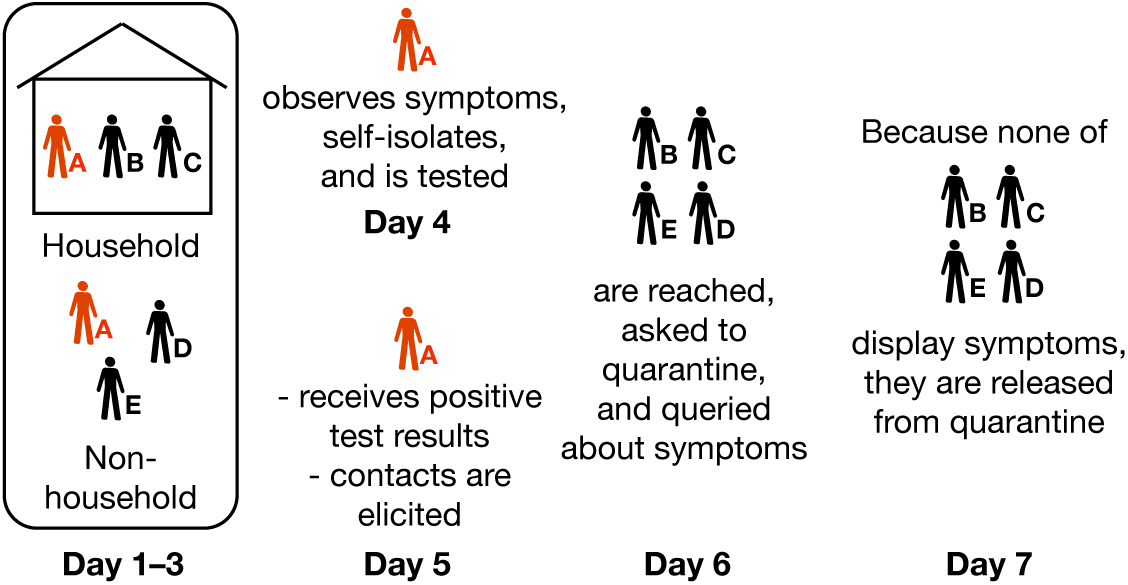
Risk-based quarantine can achieve a comparable reduction in transmission compared to quarantine-only contact tracing and requires fewer quarantine days. On day 1–3, Subject A is unknowingly infectious and exposes Subjects B–E. On day 4, A observes symptoms, self-isolates and is tested. On day 6, B–E are reached and checked for symptoms. Because none of B–E display symptoms, the cluster is released on day 8.

Because RBQ uses the symptoms of close contacts to drive decision-making, it is necessary that our model account for the observation of symptoms that are not caused by SARS-CoV-2. These may be due to another illness such as influenza or due to “worried well” behavior. We set the chance that an individual experiences “false-positive” COVID-19 symptoms to 1% per day, consistent with the higher end of estimates by Hinch et al. (2020). Because contact tracers ask the close contacts if they have experienced any symptoms since the day of exposure, we observe false-positive symptoms in about 10% of close contacts. Given that close contacts are observed on average for about a week, this rate is somewhat higher than rates of reporting of influenza-like symptoms observed by Smolinski et al. (2015) of 13–17% per season.

Of course, the effectiveness of such an RBQ system depends on both the size of the cluster and how long it is observed for. A large cluster that does not display symptoms is a much stronger indication that the index case is not highly transmissive than a small cluster that does not display symptoms. Likewise, the longer the period of observation without symptoms in the cluster, the more likely an individual in the cluster is not infectious. To illustrate this point, Figure 3 illustrates the probability that a released individual is actually infectious under RBQ. Each line represents a different cluster size, while the lines show how results vary with duration of quarantine before release. For example, if an index case has a single contact (top line of Figure 3), the probability that the contact will eventually be infected given that they do not have symptoms on the day the contact tracer contacts them is about 20%. If we instead have a cluster of size 9 (bottom line of Figure 3), the equivalent probability is 7.5%.

**Figure 3:**
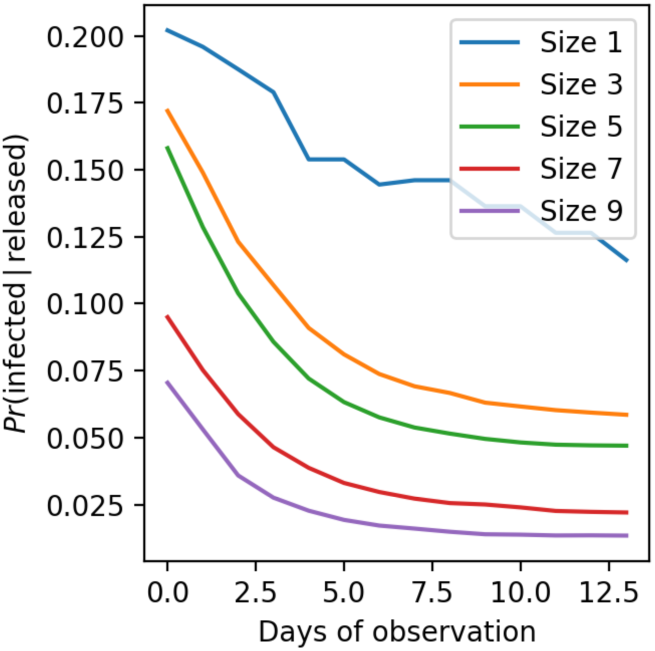
The probability that an individual becomes infected given that their entire cluster is asymptomatic by day *x*. Individuals from larger clusters are less likely to be infected when released from quarantine under RBQ.

### 3.1 More conservative extensions of risk-based quarantine

Combining RBQ with other interventions can further reduce transmission at an additional cost, either monetary or in quarantine days served.

#### Exit testing

By testing contacts who are eligible for release under baseline RBQ, some asymptomatic and pre-symptomatic cases can be detected and isolated.

#### Observing smaller clusters for longer

A consequence of the simplicity of baseline RBQ is that the risk of infection at the time of release varies dramatically by cluster size (Figure 3). Larger clusters provide more information about the infectiousness of the index case because there are more samples; furthermore, smaller clusters have a larger share of household close contacts on average. To reduce the variation in the amount of risk incurred, we can prescribe additional observation days for clusters that are below a particular size. Observing smaller clusters for a few additional days can further reduce transmission while only marginally increasing the amount of quarantine.

#### Active monitoring

Because COVID-19 symptoms are often initially very mild, symptoms can easily be missed by infected individuals, which is reflected in the observed average delay of 1.5 days between onset and self-isolation (Bi et al. 2020). We consider active monitoring, where each individual completes an app-based symptom screening each day. Digital approaches to syndromic surveillance, such as Flu Near You (Smolinski et al. 2015), have proven to be effective ways of eliciting long-term engagement with symptom surveillance systems through web or phone-based polling (Lapointe-Shaw et al. 2020). We view the use of app-based technologies as a supplement to existing contact tracing strategies, providing an additional communication platform to share resources and provide support.

We model active monitoring as reducing the isolation delay to one day for those for whom it exceeds one day. Active monitoring incurs an additional cost, in that some individuals who display false-positive COVID-19 symptoms are instructed to isolate. However, these “pseudo-isolation” days may have other beneficial effects, such as reducing the spread of other respiratory infections.

### 3.2 Comparison to other contract tracing policies

Table 2 describes our contact tracing modeling assumptions. Our model of contact tracing is intended to represent a robust, but imperfect, public health system. Because of ongoing physical distancing and masking efforts, we perform a baseline reduction in non-household contacts relative to our calibrated model; namely, the probability that a given contact event does not occur is 0.5, resulting in an average of 15.8 close contacts (SD 11.6) per individual. Reaching 80% of index cases and 80% of their contacts in one day corresponds to a highly effective, but not unrealistic, contact tracing system. We assume that contact tracers are paid $20 USD per hour. Interviewing an index case takes 30 minutes plus 15 minutes per contact, and each contact that is in quarantine, isolation or active monitoring requires 10 minutes of contact tracer time per day.

A key aspect of modeling quarantine is capturing the dropout rate, which undercuts the effectiveness of the system. We assume that the probability of dropping out of quarantine is 5% per day, although we also assume that individuals who are symptomatic or who test positive do not drop out of isolation. We also assume that this dropout probability is responsive to information about the infectiousness of the cluster; in particular, we assume that the probability of dropout is halved when an individual is informed that another individual in their cluster has developed symptoms. We discuss and perform sensitivity analysis on these assumptions in the discussion section, as they are not strongly informed by data.

We assume that, at baseline, RT-PCR tests take one day to administer. As the number varies widely in practice (Lazer et al. 2020), we perform sensitivity analysis on this quantity. Our model of RT-PCR test is based on the analysis of Kucirka et al. 2020 and presents a high false-negative rate initially that drops to 25% at 3–6 days after symptom onset.

We compare baseline RBQ to several alternative contact tracing policies (Table 3). We include a baseline without tracing at all and standard quarantine-only tracing, which provide the two extremes of our analysis. Additionally, we include test-based release policies. Single-test release tests individuals immediately when they are reached by tracers and releases individuals who test negative. Double-test is a more conservative variant; after the results of each test become available, another test is taken, until two negative tests are obtained or quarantine ends. We additionally consider several variants of RBQ, including the conservative extensions from the previous section. For longer observation times for smaller clusters, we choose four extra observation days for clusters of size eight or less as a compromise between transmission reduction and additional quarantine.

**Table 3:**
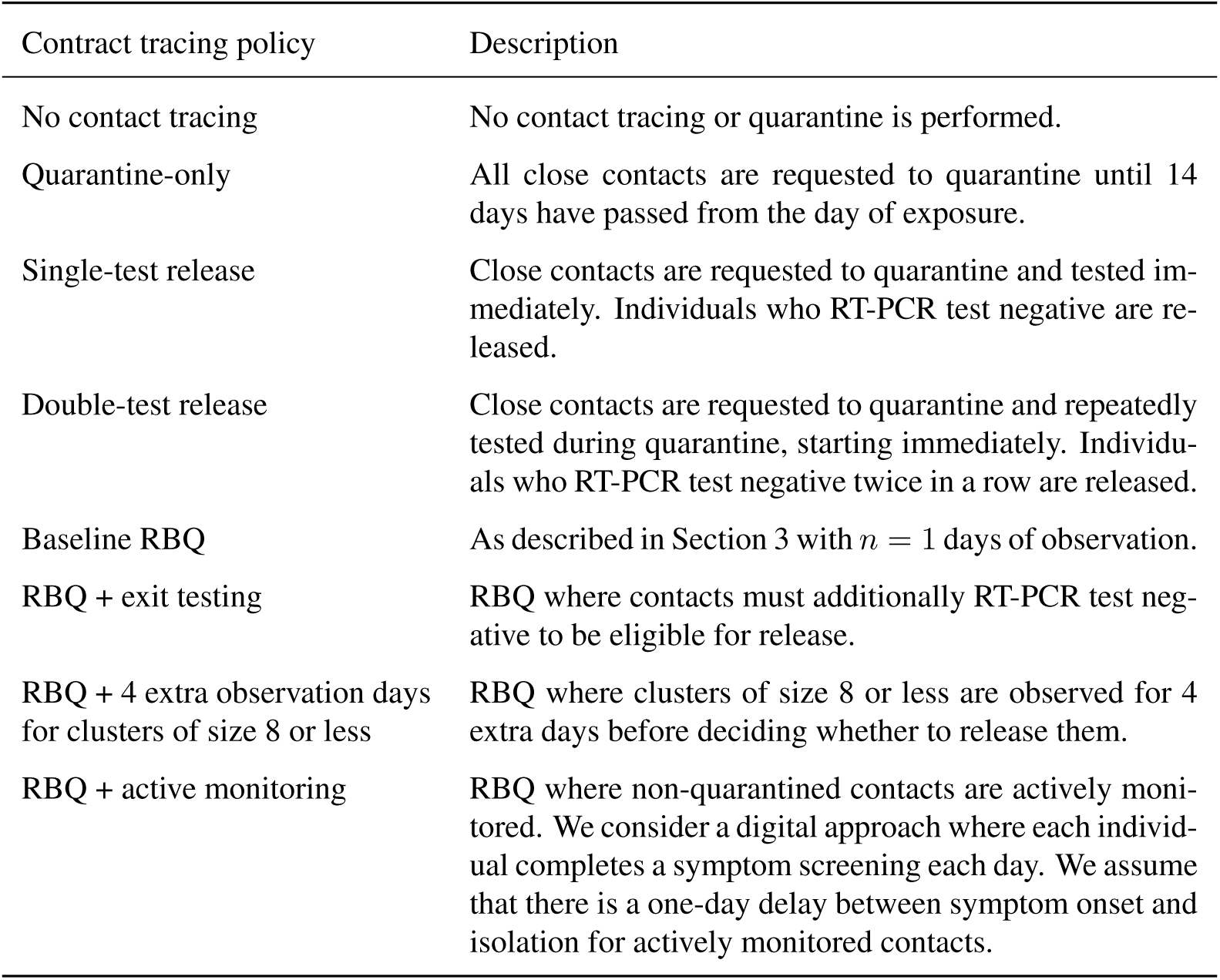
Evaluated contact tracing policies

For each policy, we simulate 100,000 index cases. The model code is available online (https://github.com/aperrault/test-trace).

## 4 Results

Our model predicts that RBQ reduces transmission more than test-based policies and costs many fewer quarantine days than standard quarantine-only tracing. RBQ results in a slightly higher level of transmission and deaths than quarantine-only tracing, but this gap can be bridged by combining the more conservative extensions to RBQ, incurring a marginal additional requirement of quarantine days and monetary cost.

Table 4 presents a comparison of alternative contact tracing and testing policies. We show the *R*_eff_ of second generation contacts, as well as the reduction in *R*_eff_ relative to a baseline where no contact tracing is present. We then show the total days of quarantine per index case among all second generation contacts. We compute deaths per 1000 index cases across second and third generation contacts. Finally, we show the cost per index case, including costs of tracing, testing and monitoring.

**Table 4:** Comparing the effectiveness of contract tracing policies

| Contract tracing policy | $R_{\text{eff}}$ of close contacts | Mean reduction in $R_{\text{eff}}$ | Quarantine days per index case | Deaths per 1000 index cases | Cost per index case |
| --- | --- | --- | --- | --- | --- |
| No contact tracing | 1.36 | 0% | 0 | 27.4 | \$0 |
| Quarantine-only | 0.926 | 31.8% | 62.1 | 22.6 | \$189 |
| RBQ | 1 | 26.1% | 36.1 | 23.8 | \$144 |
| RBQ + exit testing | 0.967 | 28.8% | 40.1 | 23.2 | \$957 |
| RBQ + 4 extra observation days for clusters of size 8 or less | 0.968 | 28.7% | 40.5 | 23.2 | \$152 |
| RBQ + active monitoring | 0.967 | 28.8% | 36.1 | 23.2 | \$208 |
| RBQ + exit testing + 4 extra observation days for clusters of size 8 or less + active monitoring | 0.926 | 31.8% | 42.6 | 22.5 | \$970 |
| Single-test release | 1.17 | 13.8% | 14.9 | 25.8 | \$1630 |
| Double-test release | 1.1 | 19.1% | 21.2 | 24.8 | \$3500 |

We find that quarantine-only contact tracing reduces transmission among second generation contacts by 31.8% on average, resulting in 17.5% fewer deaths per index case among second and third generation contacts. The cost of this reduction in transmission and deaths is 62.1 quarantine days and $189, which is spent entirely on contact tracers.

In contrast, RBQ results in a slightly higher rate of deaths, but a significant reduction of days in quarantine. In particular, baseline RBQ results in a somewhat higher *R*_eff_ of 1 (vs. 0.926 for quarantine-only) and 5% more deaths, but only requires 36.1 days of quarantine, representing a 41.9% reduction. Baseline RBQ is somewhat less expensive (24%) than quarantine-only because less contact tracer time is required to communicate with quarantined and isolated individuals.

The effectiveness of RBQ in terms of reducing deaths can be enhanced in a variety of ways. Adding exit testing, additional observation days to small clusters, or app-based active monitoring to RBQ yields similar reductions in *R*_eff_ (28.7–28.8%) and deaths per index case (23.2 per thousand, or 2.6% less than baseline RBQ). The cost of these additions varies. Exit testing has a high monetary cost of $957 per index case and adds 11% more quarantine days. The additional quarantine days are required because each individual who is eligible for release under RBQ must remain quarantined until they receive their test results. The additional observation days are inexpensive at $152 per index case and add 12% more quarantine days, slightly more than exit testing. Active app-based monitoring does not cause additional quarantine days relative to baseline RBQ and costs 44.4% more ($208 per index case). Active monitoring adds 1.6 days of monitoring-induced isolation per index case, consisting of isolation time spent by individuals who are not infected with SARS-CoV-2.

Combining all three extensions to baseline RBQ results in the same reduction in transmission as quarantine-only (31.8%) and fewer deaths (22.5 per 1000 index cases) but costs 31% fewer quarantine days (42.6 vs. 62.1). The monetary cost of the combined approach is $970 per index case. The combined approach is able to achieve the same transmission reduction with fewer quarantine days because of our assumption that quarantine adherence is improved by the RBQ protocol ^2^

We find that contact tracing policies that immediately test contacts are less effective than RBQ at preventing transmission. Single-test release, which immediately tests each traced contact and releases those who test negative, prevents just 13.8% of transmission relative to the baseline without contact tracing—less than half of the transmission reduction achieved by quarantine-only. The 5.8% reduction in deaths (to 25.8 per 1000 index cases) is similarly small. Furthermore, single-test release is 8.6 times more expensive than quarantine-only at $1630 per index case. The advantage of single-test release is the small number of quarantine days per index case. Each uninfected contact serves a single day of quarantine after they are tested, resulting in 14.9 quarantine days per index case.^3^ The initially high false positive rate of RT-PCR tests combined with the variation in time to onset of infectiousness makes it difficult to use test results to determine whether an individual should be released from quarantine.

Double-test release, where contacts who test negative twice consecutively are released from quarantine, is somewhat more effective at reducing transmission, but is much more expensive. It achieves a reduction in *R*_eff_ of 19.1% and a reduction in deaths of 9%. However, the cost is more than twice that of single-test release at $3500 per index case. Double-test release costs about 50% more quarantine days (21.2 per index case) than single-test release.

Test-based policies require rapid test turnaround to be competitive with RBQ in terms of number of quarantine days (Figure 4). Recall that our baseline scenario assumes an optimistic 24-hour time between test and results. This point is illustrated in Figure 4. Each panel in the figure shows the effect of lengthening time between test and results, first on *R*_eff_, and then on average quarantine days. Recall that our baseline scenario assumes an optimistic 24-hour time between test and results, but in practice times have typically been much longer. At a test turnaround of three days, double-test release performs similarly to RBQ, with *R*_eff_ of 1.09 (vs. 1.10), 24.7 (vs. 24.9) deaths per 1000, and 39.2 (vs. 36.3) quarantine days per index case. Double-test release is 19.2 times more expensive, however.

**Figure 4:**
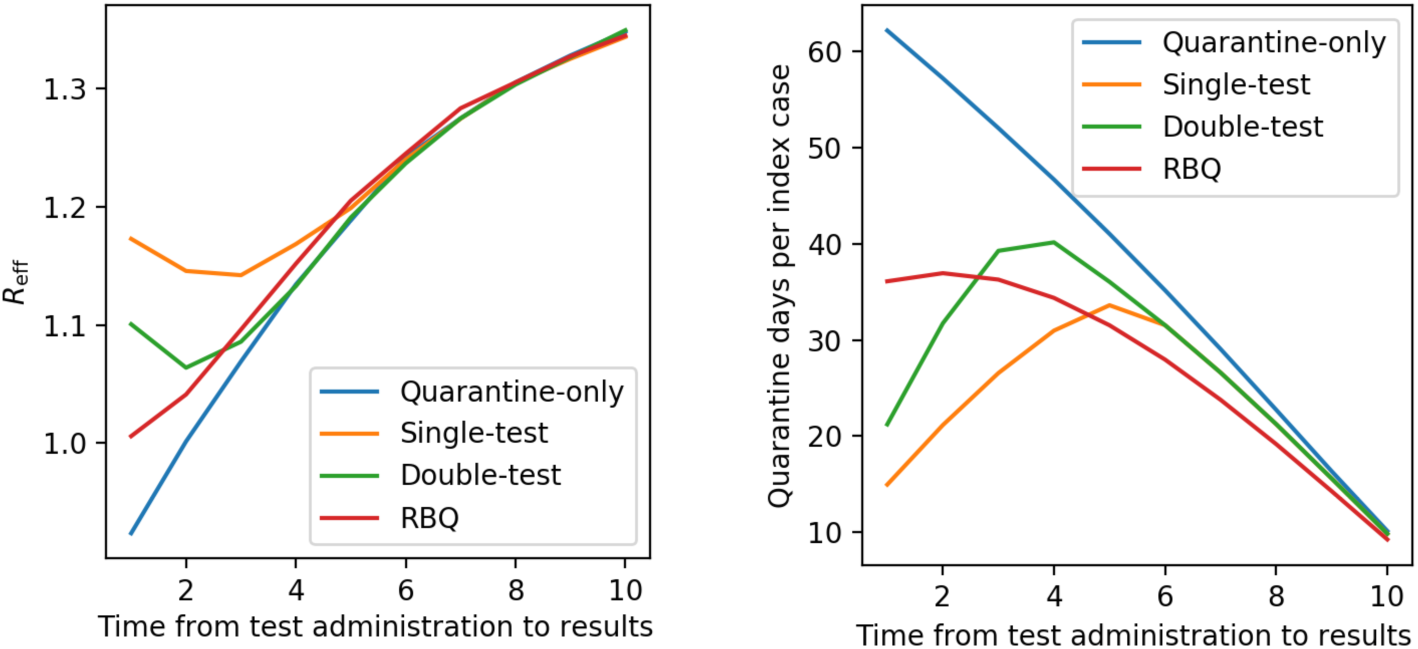
Delays in test processing reduces the effectiveness of all contact tracing policies and rapidly increases the number of quarantine days induced by test-based policies.

We find that test processing delays causes quarantine-only and RBQ to become less effective due to the need to receive a positive test result before tracing the contacts of the index case.^4^ Test-based policies initially see an increase in effectiveness as a result of test processing delays. Because it takes longer to receive a positive test result for the index case, the tests of second generation contacts are performed later, yielding results that more accurately reflect whether these individuals are infectious. The number of quarantine days for test-based release policies initially increases as individuals spend more time quarantined, waiting for test results, and then decreases because the exposure date was sufficiently long ago that quarantine ends before test results are received.

## 5 Discussion

Using a combination of data sources, we construct a branching process model of SARS-CoV-2 transmission and contact tracing. Our results are striking. We find that at baseline, with no contact tracing, each infected person causes 0.0274 deaths among their contacts, and each contact causes 1.36 infections on average. With contact tracing that requires a full 14-day quarantine, the death rate falls by 18% to 0.0226, and transmission reduces by 31.8% to 0.926 infections per contact—a substantial reduction—but the set of contacts spends an average of 62.1 days in quarantine. The long quarantine time may be quite costly and disincentivize both index cases and contacts from participating in contact tracing. On the other hand, with a baseline RBQ system, each infected person causes 5% more deaths and each contact causes 8% more transmission, but contacts spend only 36.1 days in quarantine, resulting in a 42% reduction. At a cost of 18% more quarantine days and $826 more per index case, a more conservative version of RBQ can achieve the same number of infections and deaths as standard contact tracing.

An alternative approach to reducing the number of quarantine days is to use testing to identify infected contacts and release those contacts who test negative. However, requiring a single negative test, conducted immediately, is much less effective at reducing transmission and deaths than the alternatives we consider. Transmission is reduced by 13.8% and deaths by 6%—less than half the reductions achieved by even a baseline RBQ system. Requiring two consecutive negative tests to release performs somewhat better, achieving a 19.1% reduction in transmission and a 10% reduction in deaths. While these test-based policies are effective at reducing the number of quarantine days served, with a 76% reduction for single-test release and a 65% reduction for double-test release, this advantage is quickly eliminated if there are delays in test processing. Furthermore, test-based policies are much more expensive. The combination of cost and sensitivity to test delays makes test-based policies challenging to implement in under-resourced environments.

RBQ could be extended by using properties of the contact events to inform decision-making. The more detailed the information that is available about exposure events and the more accurate a predictive model of symptoms and infection that can be constructed, the more effectively an RBQ system can perform. Despite the detail in our transmission model, we assume that contact tracers do not integrate information about the context of the contact (e.g., household vs. non-household) in their evaluation of the risk associated with the index case. By making this assumption, our findings are more robust to errors in our model of transmission. Nevertheless, in a setting where more precise models of transmission can be constructed and are available to tracers, RBQ can potentially be made considerably more effective.

Our modeling approach has significant limitations. We highlight four. First, we aggregate data from heterogeneous and possibly incompatible sources with respect to the onset and presentation of symptoms; further, there are some areas where very little relevant data are available. Our estimates of the effectiveness associated with RBQ depend critically on the fraction of individuals who develop COVID-19 symptoms after exposure and the timing of when they do so. To evaluate the effectiveness of RBQ, we need to know the probability of COVID-19 given observed symptoms and known exposure to an individual who tested positive for SARS-CoV-2 infection. The less predictive COVID-19-like symptoms are of infection, the less effective RBQ will be at reducing the amount of quarantine served. It may be desirable to observe contacts for a specific set of highly predictive symptoms, such as anosmia (Petersen and Phillips 2020). Doing so would yield higher confidence that a symptomatic individual is infected at the cost that more infected individuals would be asymptomatic under this definition.

Second, we are missing important data on the subject of adherence to quarantine and isolation. Ideally, our model would be parameterized using a distribution of the number of contacts of quarantined and isolated individuals as a function of quarantine length and motivation, i.e., information provided to the individual about the likelihood that they are sick or infectious. Unfortunately, most existing studies of quarantine adherence ask all-or-nothing questions, such as whether all conditions of quarantine were observed over the entire period, which do not inform us about the severity or timing of quarantine violations. To address this data insufficiency, we perform a sensitivity analysis around quarantine adherence assumptions in Appendix B. We find that RBQ performs better relative to quarantine-only when dropout rates are higher. This can be explained by the ability of RBQ to better exploit the lower dropout rate that we assume is a result of an individual believing they are more likely to be infected. If an individual’s belief that they are infected does not affect their quarantine dropout rate, RBQ becomes less effective at reducing transmission. Nevertheless, increases in participation in contact tracing due to shorter quarantines may still result in less transmission at the population level.

Third, the dynamics of COVID-19 transmission heterogeneity are still not well understood. The model attributes transmission heterogeneity to three causes. First, an individual can have more contacts. Second, they can have contacts that are more intense on average, e.g., because they are physical. Third, they can be more transmissive, either due to biological or behavioral factors. Our model includes all three causes, but we do not have strong evidence to support attribution of heterogeneity to each. A fourth cause that is not included in our model is variation in susceptibility, e.g., among the elderly or immunosuppressed. For RBQ, the specific balance between second, third and fourth causes does not affect our results. All of these lead to variation in the propensity of an index case to transmit the infection to their contacts, which is what RBQ exploits. Nevertheless, it is possible that transmission heterogeneity can be explained by variation in the number of contacts only. For example, if SARS-CoV-2 transmission is primarily inhalational, the definition of contact that is used by contact surveys, which requires prolonged physical proximity, may be inappropriate. This appears unlikely as studies of superspreading events to date have generally demonstrated prolonged close contact (Frieden and Lee 2020).

Fourth, our transmission model lacks detail in some areas. We assume a constant rate of asymptomatic infections across age. Age-specific asymptomatic rates would increase the effectiveness of RBQ in populations that are more likely to show symptoms and decrease it in those that are less likely. Similarly, we assume a binary, rather than continuous, model of high infectiousness that does not vary with time. The effectiveness of RBQ does not depend on this assumption as long as transmission heterogeneity remains at levels that are similar or above those we consider.

Significant obstacles must be overcome to implement RBQ in practice. System inertia is a major factor—there is a body of contact tracing best practices that do not consider differential handling based on risk. In order to understand whether the benefits are worth the transitioning costs, we recommend a two-pronged approach. First, the effectiveness of RBQ at preventing transmission can be tested empirically. In an information-rich environment, e.g., a school or university, where app-based monitoring is already in use, RBQ can be tested empirically against standard quarantine-only tracing by comparing the number of contacts of contacts who develop COVID-19 symptoms as reported by the app. Because these environments are often coercive, they are not ideal for measuring the extent to which RBQ may increase tracing participation. Thus, we secondly recommend using a survey to evaluate the extent to which RBQ might impact participation in contact tracing. We would like to measure to what extent a substantial reduction in quarantine days on average would influence the decision of individuals to provide information to contact tracing systems.

In summary, our findings suggest that approaches to contact tracing that take infection risk into account, such as RBQ, can reduce the amount of quarantine time served without increasing transmission and can do so inexpensively. By reducing the cost that quarantine imposes on individuals, they may increase participation in contact tracing, enabling less frequent and severe lockdown measures.

## Data Availability

All code used to run the simulations is available at https://github.com/aperrault/test-trace.

## Footnotes

1. The ideal duration of observation may vary depending on the amount of time it takes to test the index case and reach their contacts and the desired trade-off between transmission risk and amount of quarantine.

2. Recall that after stating that symptoms were observed in a cluster, we assume that quarantine dropout rate is halved from 5% per day to 2.5% per day.

3. Note that we do not consider introducing a delay before the first test is conducted. Doing so would reduce transmission and increase quarantine.

4. It is possible to consider contact tracing that does not require a positive test result for the index case, e.g., tracing individuals that have a highly predictive set of symptoms and an attribute that makes them more likely to be highly transmissive.

