## Appendix for "Designing Efficient Contact Tracing Through Risk-Based Quarantining"

Author affiliations:

### A Detailed Model Description

The model generates the two generations of contacts (i.e, second and third generation contacts) following an infected index case, according to the following procedure. In the following subsections, we describe the subroutines referred to here and the key assumptions made by each.

1. Generate index case. $\textsc{IndexCase} = \textsc{GenerateIndexCase}()$.
2. Generate contacts. $\textsc{ContactEvents} = \textsc{GenerateContactEvents}(\textsc{IndexCase})$.
3. Initialize $\textsc{Contacts} = []$ (empty list). For each $\textsc{ContactEvent} \in \textsc{ContactEvents}$:
   (a) Append $\textsc{TransmitInfection}(\textsc{IndexCase}, \textsc{ContactEvent})$ to $\textsc{Contacts}$.
4. Apply contact tracing to $\textsc{Contacts}$. $\textsc{Contacts} = \textsc{TraceContacts}(\textsc{IndexCase}, \textsc{Contacts})$.
5. Generate contacts of contacts. For each $j \in \textsc{Contacts}$:
   (a) $\textsc{ContactEvents}_j = \textsc{GenerateContactEvents}(j)$.
   (b) Initialize $\textsc{Contacts}_j = []$ (empty list). For each $\textsc{ContactEvent} \in \textsc{ContactEvents}_j$, append TRANSMITINFECTION(j, CONTACTEVENT) to $\textsc{Contacts}_j$.

#### A.1 GENERATEINDEXCASE

The first step in the model is to generate index cases that correspond to infected individuals who are known to the healthcare system. These individuals are assumed to have developed symptoms, self-isolated, and then tested positive. Before isolating, each index case has exposed their close contacts to SARS-CoV-2. Contact tracing, if it occurs quickly enough, can reach these contacts and instruct them to quarantine.

Pseudocode for this step can be found at the end of the section. We assume that index cases are drawn uniformly from the population distribution of the United States, reflecting the common assumption of uniform attack rate for respiratory illnesses (Verity et al. 2020). In particular, we use the distribution [18.62%, 13.12%, 39.29%, 12.94%, 16.03%] for individuals aged 0–14, 15–24, 25–54, 55–64, and 65+, respectively (Howden and Meyer 2011). We randomly match the index case to an individual of the same age range in the POLYMOD responses (Mossong et al. 2008), which are used later to determine the contacts of the index case. What remains is to determine the infection and behavior parameters of the index case using the parameters in Table 1; these will be used to determine how many days the index case was transmitting the virus and whether they were highly transmissive. We draw the time from exposure to symptoms from a log-normal distribution with log mean 1.57 days and log standard deviation of 0.65

days and the time from symptom onset to isolation from a gamma distribution with shape 1.28 days and scale 1.22 days (Bi et al. 2020). The number of transmission days for the index case is the number of days from symptom onset to self-isolation plus two (the infectious period before symptom onset). To determine whether the individual is highly transmissive, we draw a Bernoulli with success probability 10.9%.

**GENERATEINDEXCASE.** Input: nothing. Output: the parameters of an index case $i$.

1. Set the age of the index case. Draw $\textsc{Age}_i \sim \text{USP}\textsc{opulation}$.
2. Match the index case to a POLYMOD survey respondent of the same age. Draw $\text{POLYMODR}\textsc{esponse}_i \sim \text{POLYMODS}\textsc{urvey}(\textsc{age}_i)$.
3. Draw infection and behavior parameters from their distributions, including $t_{\text{incubation}}$, $t_{\text{self-isolate}}$, $I_{\text{highly transmissive}}$. Because $i$ is an index case, we fix $I_{\text{self-isolate}} = 1$ and $I_{\text{symptoms}} = 1$, $t_{\text{exposure}} = 0$, and $I_{\text{infected}} = 1$.
4. A critical derived quantity for contact modeling is the number of transmission days, the number of days that an infected individual is transmitting the virus. For index cases, $n_{\text{transmission days}} = t_{\text{self-isolate}} - t_{\text{incubation}} + \text{\# of days infectious before symptoms}$.
5. Return $i$.

#### A.2 GENERATECONTACTEVENTS

The second step in the model is to determine the contact events that occur during the infectious period of the index case. The POLYMOD survey response the individual is matched to consists of a single day of contacts, but we need to model the contacts for each transmission day of the index case. To do this, we assume that these contacts occur on each day of the transmission period. If a POLYMOD response contact is marked as daily, it is assumed to correspond to the same individual throughout the transmission period. Non-daily contacts are assumed to correspond to contacts with different individuals.

To represent the reduction in contacts due to pandemic responses such as physical distancing, we assume that each instance of a non-household contact has a 50% chance to not occur. Daily contacts will occur on fewer days during the transmission period, but it is unlikely that contact with that individual will be eliminated entirely (and less likely the longer the transmission period). Because non-daily contacts only have a single instance, each has a 50% chance to not occur.

**GENERATECONTACTEVENTS.** Input: an infected individual $i$. Output: the contacts of that individual during his/her infectious period and the days each individual was contacted on.

1. Initialize $\textsc{ContactEvents}_i = []$ (an empty list).
2. For each $\textsc{SurveyContact} \in \text{POLYMOD}\textsc{Response}_i$:
   (a) To apply the overall reduction in contacts, we draw a Bernoulli random variable to indicate for each transmission day that represents whether this contact occurs.

$$\textsc{ReductionMask}_{\text{day}}(\textsc{SurveyContact}) = \text{Bernoulli}(\text{Contact reduction }\%). \quad (4)$$

   If SURVEYCONTACT is a household contact, there is no reduction: $\textsc{ReductionMask}_{\text{day}}(\textsc{SurveyContact}) = 0$.

   (b) If SURVEYCONTACT is a daily contact, and $\min(\textsc{ReductionMask}(\textsc{SurveyContact})) = 0$, then the contact occurred on the days that $\textsc{ReductionMask}(\textsc{SurveyContact}) = 0$. Append SURVEYCONTACT augemented with the days the contact occurred to $\textsc{ContactEvents}_i$.

   (c) If SURVEYCONTACT is a non-daily contact, for each day where $\textsc{ReductionMask}(\textsc{SurveyContact}) = 0$, append SURVEYCONTACT and the day of occurrence to $\textsc{ContactEvents}_i$.
3. Return CONTACTEVENTS.

#### A.3 TRANSMITINFECTION

The third step of the model is to determine to which contacts the virus is transmitted and the parameters of these infections. The transmission probability is calculated according to Equation 3; there is a base transmission chance of 3.56% for a household contact and 0.669% for a non-household contact, which is multiplied by 24.4 if the index case is highly transmissive, by 5.21 if the contact is physical, by 0.36 if the index case is asymptomatic, and 0.63 if the index case is pre-symptomatic (the properties of each contact are provided by autocitemossong2008social). We assume that the number of days of contact does not affect the probability of transmission (i.e., transmission, if it occurs, takes place on a random day of contact). The last day of exposure is also recorded, as it is used to determine quarantine length.

The infection and behavior distributions in Table 1 determine the parameters of the infection for each infected contact. The age of the contact is taken directly from the POLYMOD survey response. The contact develops symptoms with probability 0.8 and, if they do so, self-isolates with probability 0.9. Recall that among individuals who are not index cases, none seeks testing unless the contact tracing model prescribes it. The incubation time, time to self-isolation, and whether the contact is highly transmissive are determined as they were for an index case in Section A.1.

**TRANSMITINFECTION.** Input: an infectious individual $i$ and a CONTACTEVENT. Output: either an infected individual $j$ or an exposed, non-infected, individual $j$.

1. Calculate the probability that $j$ becomes infected. $t_{\text{exposure}}$ drawn uniformly from the days that CONTACTEVENT occurred. Let BASE $=$ BASE$_{\text{household}}$ if CONTACTEVENT is a household contact and base $=$ base$_{\text{ext}}$ otherwise. Let MULT$_{\text{phys}}$ be a multiplier that increases the probability of infection if the contact is physical and is 1 otherwise, and let MULT$_{\text{highly transmissive}}$ be a multiplier that increases the probability of infection if $i$ is highly transmissive. Let MULT$_{\text{symptoms}}$ be an infectiousness multiplier caused by the presence or absence of symptoms in individual $i$ on $t_{\text{exposure}}$.

$$Pr(i \text{ infects } j) = \min(\text{BASE} * \text{MULT}_{\text{phys}} * \text{MULT}_{\text{highly transmissive}} * \text{MULT}_{\text{symptoms}}, 1) \quad (5)$$

2. Create a new individual $j$. AGE$_j$ is copied from CONTACTEVENT. If Bernoulli($Pr(i \text{ infects } j)$), set $I_{\text{infected}} = 1$. Otherwise, set $I_{\text{infected}} = 0$. The remaining infection and behavior parameters are drawn from their distributions, including $t_{\text{incubation}}$, $I_{\text{symptoms}}$, $I_{\text{self-isolate}}$, $t_{\text{self-isolate}}$, $I_{\text{highly transmissive}}$. $t_{\text{last exposure}}$ is the latest day of contact between $i$ and $j$ and is used to calculate quarantine lengths.

3. Return $j$.

#### A.4 TRACECONTACTS

In the fourth step of the model, we modify the behavior of contacts according to the contact tracing policy. The index case is reachable by tracers with probability 0.8, i.e., they agree to provide their contacts in an interview. The contact tracer then attempts to reach each second generation contact, which is successful with probability 0.8 (independent for each contact).

What happens in the case of successfully reaching a contact varies by contact tracing policy. In quarantine-only, the contacts are requested to quarantine for 14 days from the last day of contact. In $k$-test release, each contact is quarantined and tested until they either test negative $k$ consecutive times, in which case they are released, or until 14 days from the last day of contact. RBQ is described in Section 3. We assume that, if a contact develops symptoms before or during quarantine, they are isolated for 10 days from the day of symptom onset (this may be shorter or longer than the 14-day quarantine period).

After the requested quarantine period is determined, the behavior of the contact is simulated on a daily basis. They drop out of quarantine with a base probability of 5% per day. If they test positive or develops symptoms, the probability drops to 0%. If they test negative, it doubles to 10%. In the case of RBQ, we assume that the probability of dropout halves to 2.5% per day if the contact is informed that symptoms developed among others who were exposed to the same index case. The key results of the simulation are the number of transmission and quarantine days for each contact, the cost of quarantine and testing, and whether each contact died. For the last, we use the infection fatality rates of (Levin et al. 2020): 0.01% for ages 0–24, 0.13% for 25–54, 0.7% for 55–64, and 6.8% for 65+.

**TRACECONTACTS.** Input: an infectious individual $i$ and his/her CONTACTS. Output: an update to each contact that adds the number of transmission, quarantine and isolation days.

1. Are tracers able to reach $i$? If Bernoulli(prob. index case reached) is 0, we are unable to reach $i$ and no tracing occurs and the parameters of CONTACTS are not updated.
2. For each $j \in$ CONTACTS, are tracers able to reach that contact? Sample Bernoulli(prob. contact reached). Let REACHEDCONTACTS be the set of contacts that tracers successfully reach.
3. Apply a contact tracing policy to modify the behavior of each reached contact. For example, quarantine-only tracing sets $t_{\text{quarantine start}}$ for each reached contact equal to the sum of the index case's $t_{\text{self-isolate}}$ plus the time to receive test results and the time to trace contacts (simulating the time required to test the index case and trace his/her contacts), and sets $t_{\text{quarantine end}}$ to $t_{\text{last exposure}}$ plus quarantine length.
4. For each $j \in$ CONTACTS simulate his/her behavior during the quarantine and infectious period, recording the number of transmission, quarantine and isolation days. This process includes simulating quarantine dropout, changing the release date due to the development of symptoms, monitoring and false-positive isolation.
5. Return CONTACTS.

#### A.5 Transmission to Contacts of Contacts

The fifth and final step of the model is to calculate the amount of transmission from the contacts of index cases to their contacts and the number of deaths that occur. We take the same approach used to generate contacts for index cases (i.e., match each contact to a survey response in POLYMOD with the same age as the contact and use GENERATECONTACTEVENTS). We assume (pessimistically) that there is no triadic closure, i.e., the contacts of the contact do not overlap with the contacts of the index case. Applying TRANSMITINFECTION determines which of

Table 5: Doubled quarantine dropout rate

| Contract tracing policy | $R_{\text{eff}}$ of close contacts | Mean reduction in $R_{\text{eff}}$ | Quarantine days per index case | Deaths per 1000 index cases | Cost per index case |
|---|---|---|---|---|---|
| No contact tracing | 1.36 | 0% | 0 | 27.4 | $76 |
| Quarantine-only | 0.968 | 28.7% | 50.2 | 23.2 | $168 |
| RBQ | 1.03 | 24.2% | 30.9 | 24.2 | $135 |
| RBQ + exit testing | 0.994 | 26.7% | 35.1 | 23.6 | $969 |
| RBQ + 4 extra observation days for clusters of size 8 or less | 0.995 | 26.7% | 35.5 | 23.7 | $143 |
| RBQ + active monitoring | 0.965 | 28.9% | 36.1 | 23.3 | $208 |
| RBQ + exit testing + 4 extra observation days for clusters of size 8 or less + active monitoring | 0.959 | 29.3% | 37.8 | 23 | $991 |
| Single-test release | 1.18 | 13.3% | 14.2 | 25.9 | $1470 |
| Double-test release | 1.11 | 18.5% | 19.1 | 24.9 | $3050 |

these contacts become infected. The end result is that we can compare contact tracing policies with respect to their impact on the number of infections and deaths in the second and third generation and the cost incurred in the second. (Because there is no tracing in the third generation, no costs are incurred.)

### B Sensitivity Analysis

To test the sensitivity of the model to quarantine dropout rate assumptions, we reproduce Table 4 when quarantine dropout rate is doubled (Table 5), when knowledge of symptoms in the same cluster is 50% more effective at preventing dropout (Table 6), and when both adjustments are made simultaneously (Table 7).

Increases in quarantine dropout rate hurt all policies, but impact quarantine-only the most. Increasing the impact of knowledge of symptoms in the same cluster makes RBQ more effective, but the impact is marginal.

Table 6: 75% dropout reduction for RBQ after notification

| Contract tracing policy | $R_{\text{eff}}$ of close contacts | Mean reduction in $R_{\text{eff}}$ | Quarantine days per index case | Deaths per 1000 index cases | Cost per index case |
|---|---|---|---|---|---|
| No contact tracing | 1.36 | 0% | 0 | 27.4 | $76 |
| Quarantine-only | 0.926 | 31.8% | 62.1 | 22.6 | $189 |
| RBQ | 0.996 | 26.6% | 37.2 | 23.8 | $146 |
| RBQ + exit testing | 0.961 | 29.2% | 41.3 | 23.1 | $959 |
| RBQ + 4 extra observation days for clusters of size 8 or less | 0.96 | 29.2% | 41.0 | 23.1 | $152 |
| RBQ + active monitoring | 0.957 | 29.5% | 37.3 | 22.9 | $210 |
| RBQ + exit testing + 4 extra observation days for clusters of size 8 or less + active monitoring | 0.92 | 32.2% | 43.1 | 22.5 | $971 |
| Single-test release | 1.17 | 13.8% | 14.9 | 25.8 | $1630 |
| Double-test release | 1.1 | 19.1% | 21.2 | 24.8 | $3500 |

Table 7: Doubled dropout rate and 75% dropout reduction for RBQ after notifcation

| Contract tracing policy | $R_{\text{eff}}$ of close contacts | Mean reduction in $R_{\text{eff}}$ | Quarantine days per index case | Deaths per 1000 index cases | Cost per index case |
|---|---|---|---|---|---|
| No contact tracing | 1.36 | 0% | 0 | 27.4 | $76 |
| Quarantine-only | 0.968 | 28.7% | 50.2 | 23.2 | $168 |
| RBQ | 1.02 | 25.1% | 32.8 | 24.1 | $138 |
| RBQ + exit testing | 0.981 | 27.7% | 36.9 | 23.3 | $972 |
| RBQ + 4 extra observation days for clusters of size 8 or less | 0.99 | 27.1% | 36.2 | 23.6 | $144 |
| RBQ + active monitoring | 0.959 | 29.4% | 37.2 | 23.1 | $210 |
| RBQ + exit testing + 4 extra observation days for clusters of size 8 or less + active monitoring | 0.946 | 30.3% | 38.6 | 22.9 | $993 |
| Single-test release | 1.18 | 13.3% | 14.2 | 25.9 | $1470 |
| Double-test release | 1.11 | 18.5% | 19.1 | 24.9 | $3050 |
